# An Evidenced-Based Prior for Estimating the Treatment Effect of Phase III Randomized Trials in Oncology

**DOI:** 10.1101/2024.04.01.24305158

**Authors:** Alexander D. Sherry, Pavlos Msaouel, Gabrielle S. Kupferman, Timothy A. Lin, Joseph Abi Jaoude, Ramez Kouzy, Zachary R. McCaw, Ethan B. Ludmir, Erik van Zwet

**Author notes:** **Correspondence:** Dr. Erik van Zwet, Biomedical Data Sciences, Leiden University Medical Center, Einthovenweg 20, 2333 ZC Leiden, The Netherlands. **Data availability:** The underlying dataset, code for the development of the prior, and code for computing posterior probability are provided in the Supplement. A freely available webpage has been created for users to compute the probabilities at a given level of hazard ratio based on the trial’s summary statistics: https://alexandersherry.shinyapps.io/shinyapp/. **Funding:** Supported in part by Cancer Center Support (Core) grant P30CA016672 from the National Cancer Institute to The University of Texas MD Anderson Cancer Center and by the Sabin Family Fellowship Foundation (E.B.L. and P.M.). **Contributions**: A.D.S., P.M., Z.R.M., E.B.L., and E.V.Z. conceptualized and designed the study. A.D.S., G.S.K., T.A.L., J.A.J., R.K., and E.B.L. curated the data. A.D.S., Z.R.M., and E.V.Z. did the formal analyses. E.B.L. acquired funding. P.M., E.B.L., and E.V.Z supervised the study and provided study resources. A.D.S. and E.V.Z. prepared the visualizations. All authors contributed to the interpretation of the data. A.D.S. wrote the first draft, and all authors reviewed and revised the manuscript. All authors had full access to all data in the study and accept responsibility to submit for publication.

## Abstract

**Purpose:** The primary results of phase III oncology trials may be challenging to interpret, given that such results are generally based on meeting *P*-value thresholds. The probability of whether a treatment is beneficial, although a more intuitive summary of the results, is not provided by most trials. In this study, we developed and released a user-friendly tool that calculates the probability that a treatment studied in a phase III oncology trial is beneficial using published summary statistics.

**Methods:** We curated the primary time-to-event outcomes of 415 phase III, superiority design, therapeutic randomized controlled trials of oncologic treatments enrolling 338,600 patients and published between 2004 and 2020. A phase III oncology-specific prior probability distribution for the treatment effect was developed based on an estimated three-component zero-mean mixture distribution of the observed z-scores. Using this prior, we computed the probability of any benefit (hazard ratio < 1) and the probability of clinically meaningful benefit (hazard ratio < 0.8) for each trial. The distribution of signal-to-noise ratios of phase III oncology trials was compared with that of 23,551 randomized trials from the Cochrane Database of Systematic Reviews.

**Results:** The signal-to-noise ratios of phase III oncology trials tended to be much larger than randomized trials from the Cochrane database. Still, the median power of phase III oncology trials was only 49% (IQR, 14% to 95%), and the power was less than 80% in 65% of trials. Using the developed phase III, oncology-specific prior, only 53% of trials claiming superiority (114 of 216) had a ≥ 90% probability of providing clinically meaningful benefits. Conversely, the probability that the experimental arm was superior to the control arm (HR < 1) exceeded 90% in 17% of trials interpreted as having no benefit (34 of 199).

**Conclusion:** By enabling computation of contextual probabilities for the treatment effect from summary statistics, our robust, highly practical tool, now posted on a user-friendly webpage, can aid the wider oncology community in the interpretation of phase III trials.

## INTRODUCTION

The interpretation of modern phase III randomized trials in oncology is a considerable challenge.^1^ The standard approach for estimating comparative survival advantages is to compute hazard ratios (HRs) and their 95% CIs, with most trials declaring superiority of an experimental intervention based on *P*-value thresholds.^2^ However, 95% CIs and *P* values are widely misinterpreted. 95% CI are often misunderstood as having 95% probability of containing the true effect, and *P*-values are often mistaken as the probability of no difference.^3–5^ *P*-value thresholds may lead to both overestimation and underestimation of effects, particularly in scenarios where power is lower than planned.^6–9^ For example, a significant *P* value (e.g., P < 0.05) in a trial designed with 80% power does not imply an 80% probability that the experimental treatment was beneficial. Therefore, novel tools to improve the interpretation of primary outcomes in oncology trials are sorely needed. Some have proposed directly computing the probability of benefit (e.g., HR < 1) using Bayesian approaches, because the probability of whether an intervention is helpful or harmful is more intuitive to oncologists and patients than interpreting *P* values.^10–13^ However, such calculations require specification of prior knowledge, which may appear controversial due to apparent subjectivity even when guided by domain expertise.^11,14,15^

There is a considerable need for a straightforward, data-driven approach to estimating the probability of benefit in phase III trials, including in the absence of individual-level patient data, which are often difficult for clinicians to access. Here, we propose a user-friendly and evidence-based solution for estimating the probability of whether new oncology treatments tested in phase III trials are effective using standard trial-level summary statistics. The purpose of the present study was to develop an informative, oncology-specific default prior, derived from the distribution of the z-scores obtained from 415 contemporary phase III oncology trials. This default prior can be used by practicing oncologists to interpret historical, current, and future phase III oncology trials.

## METHODS

Institutional review board approval for this study was not needed due to the public availability of the trial data. Data for development of the prior were obtained from the primary results of phase III trials registered in ClinicalTrials.gov and identified using previously reported search criteria.^16^ Phase III, two-arm, superiority-design, oncology trials testing therapeutic anti-cancer strategies using a time-to-event primary endpoint were included (**Figure 1**). Primary endpoint summary statistics (HRs and 95% CIs) were recorded. The control arm was taken as the reference for all comparisons, such that HR < 1 always favored the experimental arm and in cases where the experimental arm was set as the reference, reciprocals for the HR were used. For trials with multiple co-primary endpoints, the time-to-event primary endpoint with a reported 95% CI was used. If 95% CIs were used for all time-to-event co-primary endpoints, overall survival was used due to its intrinsic value and potential advantages compared with surrogate endpoints.^2,17,18^

**Figure 1.**
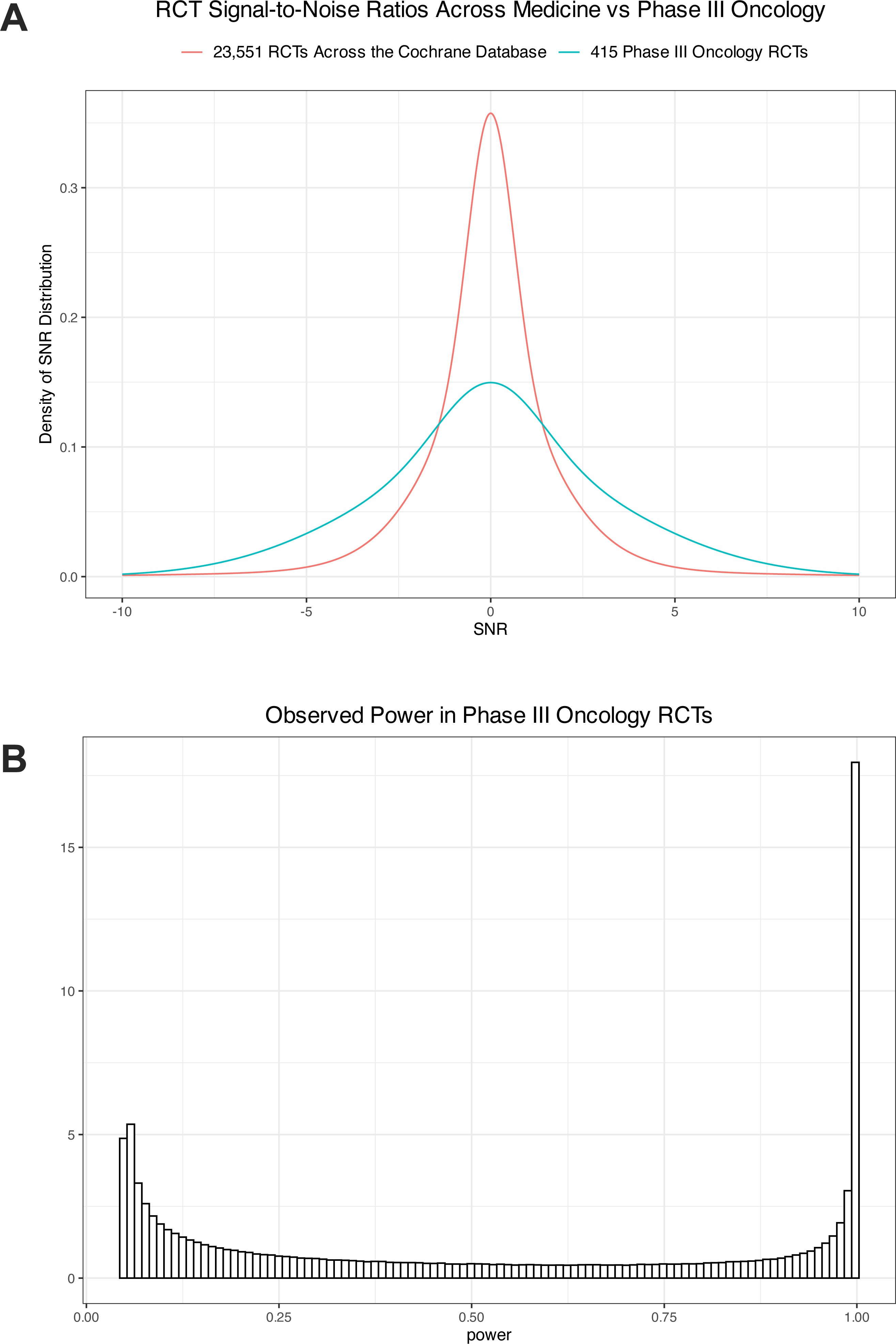
Distribution of the signal-to-noise ratio (SNR) and power of the primary outcomes of phase III randomized clinical trials (RCTS) in oncology. (A) The SNR in phase III oncology trials tended to be much larger than that of randomized trials in the Cochrane database. (B) Estimated distribution of power against the true effect in phase III oncology RCTs.

In previous work, an informative prior for addressing treatment effect exaggeration was developed using 23,551 randomized trials in the Cochrane Database of Systematic Reviews.^9,19–22^ In the present study, we applied this methodology for the creation of a phase III, oncology-specific prior distribution of the treatment effect.^19–22^ In brief, the z-score for each RCT was computed as the point estimate for the log hazard ratio (log HR) divided by the standard error.^21^ Recall that the P value is less than 0.05 when the z-score is greater than 1.96 or less than −1.96. Similar to the Cochrane-based prior distribution, we then fit a zero-mean three-component normal mixture to the z-scores.^22^ We used three components due to the smaller number of phase III oncology trials compared with the four component mixture used for the Cochrane-derived prior distribution. We chose a zero-mean mixture in order to set the prior probability of any benefit to be 50%-50%. In this way, neither treatment arm was favored in the prior distribution. While the z-score is the ratio of the estimated treatment effect to the standard error, the signal-to-noise ratio (SNR) is defined as the ratio of the *true* treatment effect to the standard error. Because the z-score represents the sum of the SNR and a standard normal error term, the “deconvolution trick” was applied to obtain the distribution of the SNR by subtracting 1 from the variances of each mixture component.^20^ It is quite remarkable that it is possible to obtain the distribution of the SNR because the true effect cannot be observed. The distribution of the power was then obtained as a transformation of the SNR.^9^ Note that this is the power against the (unobserved) true effect, and not the so-called “post hoc” power, or the power against the effect that was assumed in the sample size calculation.^23^ Lastly, the prior for the treatment effect was obtained by scaling the distribution of the SNR by the standard error.^19^ The underlying dataset and code for the development of the prior are provided in the **Supplement**. A webpage has been created for users to compute hypothesis probabilities at a given level of HR based on the trial’s summary statistics (https://alexandersherry.shinyapps.io/shinyapp/).

Based on guidelines from the American Society of Clinical Oncology, the minimum clinically important difference (MCID) was defined by HR < 0.8.^24–27^ The probability of any benefit was defined by HR < 1. The probability for both hypotheses was computed for each trial. Analyses and plots were completed in R v.4.3.2 (Vienna, Austria) and Prism v10 (La Jolla, CA).^28^

## RESULTS

After screening 785 phase III randomized trials from ClinicalTrials.gov, we included 415 two-arm, superiority-design, interventional, therapeutic, time-to-event, phase III trials (**Figure S1**). Publication dates of the primary endpoint ranged from 2004 to 2020, with a total of 338,600 patients enrolled. Most trials studied metastatic solid tumors (n=263, 63%), and most utilized surrogate primary endpoints (n=250, 60%) (**Table 1**).

Superiority was claimed for the experimental arm in 216 trials (52%) and was not claimed in 191 trials (46%); in 8 trials, inferiority of the experimental arm was claimed (2%).

The absolute z-scores of phase III oncology trials tended to be much larger than those of 23,551 RCTs from the Cochrane database (*P* < 0.0001, Mann-Whitney test) (**Figure S2**). This implies that both the SNR and the power of phase III oncology RCTs also tends to be much larger than that of general RCTs (**Figure 1A**). A zero-mean mixture of 3 normal distributions provided a reasonable fit to the z-scores of the phase III trials (**Figure S3**). The proportions and variances of each subcomponent are reported in **Table S1**. We derived a zero-mean mixture of 3 normal distributions for the SNR by subtracting 1 from each of the variances (**Table S1**). From the distribution of the SNR, we derived the distribution of the power. We find that the median power of phase III oncology RCTs was only 49% (IQR, 14% to 95%), with an average power of 52% (**Figure 1B**). An estimated 65% of trials had power less than 80%, with 71% of trials < 90%. The power was > 95% in an estimated 25% of trials. As previously reported, the power of RCTs from the Cochrane database tended to be even lower (median power: 13%, with 78% of trials < 80% power).^9^ The SNR distribution was scaled by the observed standard error to derive the prior for the treatment effect in a particular trial (**Figure S4**). Examples of resulting priors are plotted in **Figure S5.**

The probabilities of benefit (HR < 1) exceeded 90% for all 216 trials that claimed superiority (**Figure 2**). However, only 53% of trials with superiority claims (114 of 216) had ≥ 90% probability of achieving the MCID (**Table 2**). Conversely, for the 199 trials that did not claim superiority, the median probability that the experimental arm more effective than the control arm (HR < 1) was 63% (IQR, 32% to 86%) (**Figure 2**). In 17% of trials that did not claim superiority (34 of 199), the probability that HR was less than 1 exceeded 90% (**Table 2**). Consistent with the differences in the SNR between phase III oncology RCTs and RCTs from the Cochrane database, posterior probabilities computed by the Cochrane database prior appeared to be over-corrected compared with the phase III oncology-specific prior (**Figure 3**).

**Figure 2.**
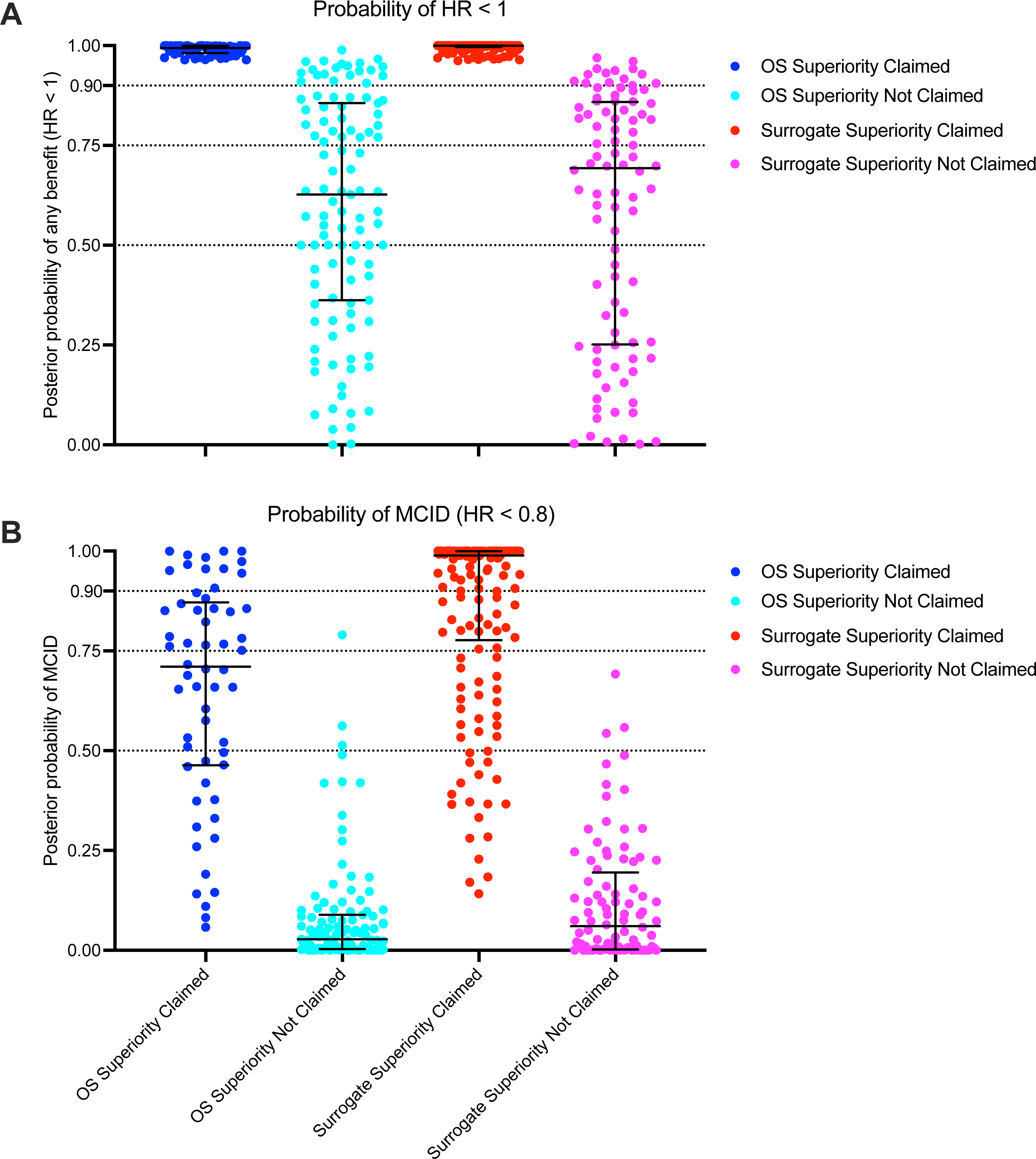
Posterior probabilities of primary endpoints of 415 phase III trials computed using the phase III oncology prior. Probabilities are grouped according to endpoint type (overall survival [OS] or surrogate survival) and the trial result interpretation (claim for superiority or not). (A) Probability that the hazard ratio (HR) is less than 1 in favor of the experimental arm. (B) Probability that the experimental arm shows superiority according to the minimum clinically important difference (MCID), defined as HR < 0.8.

**Figure 3.**
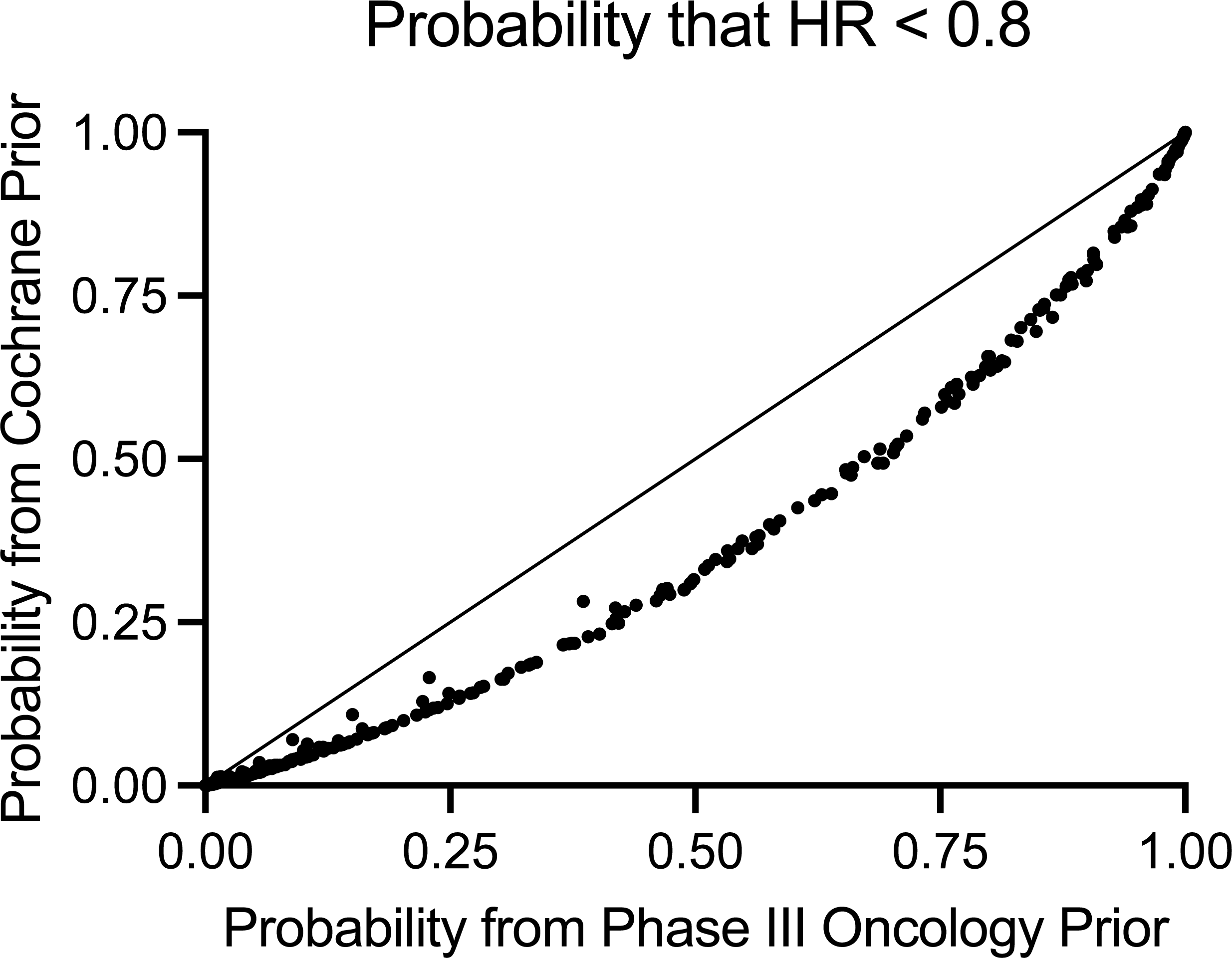
Comparison of posterior probabilities computed by the phase III oncology-specific prior versus the prior from the Cochrane database of RCTs for the minimum clinically important difference (hazard ratio < 0.8).

## DISCUSSION

Using the distribution of the z-scores of the primary endpoints of 415 phase III oncology randomized controlled trials, the present study is the first to compute an evidenced-based default prior specifically designed to estimate the effects of phase III oncology trials. This prior has been deployed in a standalone webpage application that allows users to input the summary Cox regression statistics of a phase III trial and compute the posterior probabilities of benefit at any level of hazard ratio. By providing oncologists with the means to directly compute probabilities of interest, the present study provides a robust, highly practical tool to immediately enhance the interpretation of phase III trials throughout the wider oncology community.

Consistent with previous work, we found that the actual power of most phase III trials is low relative to the power specified during trial design.^9,29^ Lower power increases the risk of false negative findings. When an underpowered trial does reach “statistical significance”, the effect is usually overestimated and leads to replication failure. Directly computing the probability of benefit using our phase III prior provides a more intuitive method of understanding and interpreting the uncertainty associated with underpowered trials. The consequences of relying on *P*-value thresholds in underpowered trials are directly manifested in our finding that the experimental arms of 17% of trials interpreted as negative or inconclusive had greater than 90% probability of superiority to the control arm.

Previously, an informative prior, based on thousands of clinical trials in the Cochrane database, was proposed.^19–22^ However, we found that the distribution of SNRs of RCTs in the Cochrane database tend to be lower than that of phase III oncology trials. Phase III oncology trials are often larger than general medical RCTs, leading to more power. The use of a phase III-specific, oncology-specific prior improves the robustness of computed posterior probabilities by reducing the risk of over-correction, as suggested by our comparison of posterior probabilities for the MCID. However, low SNR were observed even for phase III oncology trials, which can lead to upward bias in the estimate of HR.^22^ Application of the phase III oncology prior distribution of treatment effect to trials, especially those with low SNR, may partially reduce this bias, and posterior mean estimates for HR are computed as part of the provided webpage. Thus, the present study represents a considerable advance and adds strong and unique value to the interpretative tools available to oncologists.

To illustrate the potential value of estimating the probability of benefit, consider the results of two example phase III trials, the GEMPAX trial and CALGB 30610 (Alliance) / RTOG 0538, neither of which was included in the development of the phase III prior as both trials were recently published.^30,31^ The GEMPAX trial compared second-line gemcitabine with paclitaxel versus gemcitabine alone for patients with metastatic pancreatic ductal adenocarcinoma.^30^ GEMPAX showed an improvement in progression-free survival (HR 0.64, 95% CI 0.47 to 0.89) and overall response rate, but interpreted the primary endpoint of overall survival as “statistically negative” (HR 0.87, 95% CI 0.63 to 1.20) on the basis of a large *P* value (0.41). Importantly, large *P* values do not support the null hypothesis, and in fact provide little information.^4^ Using the phase III prior developed in the present study, the probability that gemcitabine plus paclitaxel is associated with better overall survival (HR < 1) than gemcitabine alone is 78%, and is similar to the 75% probability that there is no clinically meaningful difference (0.8 < HR < 1.25) between the two treatments. This highlights that the overall survival results lacked the power and precision to make reliable assertions regarding treatment efficacy or lack thereof. Conversely, the CALGB 30610 (Alliance) / RTOG 0538 RCT noted a HR of 0.94 (95% CI 0.76 to 1.17, *P* = 0.594) for OS among patients with limited-stage small-cell lung cancer who underwent one-daily radiation compared with those who underwent standard-of-care twice daily radiation.^31^ Using our phase III prior we can see that the probability that the two treatments yielded no clinically meaningful difference (0.8 < HR < 1.25) was 95%. Therefore, although both RCTs yielded a similar p-value, only RTOG 0538 had adequate power to conclusively determine no clinically meaningful difference. Notably, the standard error of the natural logarithm of HR for RTOG 0538, 0.11, was less than that of GEMPAX, 0.16. A trial with a smaller standard error is expected to have a narrower prior for the treatment effect, because treatment effect and standard error are inversely related. For example, to obtain 80% power in a trial, the treatment effect must be 2.8 times the standard error. Accordingly, in this example, GEMPAX was powered for a larger treatment effect (HR of 0.625) compared with RTOG 0538 (HR of 0.77).^30,31^ Trials expecting small treatment effects and choosing large sample sizes to reduce the standard error will result in narrower priors for the treatment effect, because the trialists themselves suspected smaller treatment effects. This distinction, made manifest in the computation of posterior probabilities by our prior distribution, highlights an example of the importance of comprehensively evaluating RCT results using our provided webtool. Importantly, probabilities of benefit do not represent rules for decision-making.^1^ Inference, obtained from data such as this study, must be applied by the oncologist to each clinical scenario in light of the risks, alternatives, patient characteristics, and patient values.^26,32^ Nonetheless, the additional information provided by posterior computation can facilitate a more informed and data-driven approach to clinical care.

There are some important limitations to consider in the present study. First, we assumed that the Cox regressions that formed the basis of the primary analysis of each phase III trial met their underlying statistical assumptions, including proportional hazards; however, this may not have always been the case.^33,34^ In general, we suggest that trials should report summary statistics that are interpretable and make as few assumptions as possible.^35^ Second, there may have been underlying variation in the approach of each trial towards computing the primary endpoint; Cox models fit with prognostic covariates are likely more precise than univariable models and this may have influenced the resultant SNRs that were used to create the prior.^36^ Third, phase III trials that were used to create the prior were identified from ClinicalTrials.gov, which may not reflect older trials or global trials. Fourth, phase III trials that were not published due to non-significant results, which was estimated in one study to be as high as 7% of trials, may have resulted in a file drawer effect and influenced our treatment effect distribution, as only published studies were included.^37^ Fifth, our prior was specifically fit to the primary endpoints of phase III trials, which is both a strength and limitation. In the present study, distribution of SNRs of phase III oncology trials appeared meaningfully different from the distribution of SNRs from general medical RCTs in the Cochrane database, as well as subsequent posterior probability estimations, thus establishing the need for a separate, phase III oncology specific prior. Consequently, other trials or endpoints, such as phase II trials or even secondary endpoints of phase III trials, are expected to have lower SNRs. Our prior is not appropriate for these cases because they lack exchangeability with the trials included in our analysis. Similarly, the prior is not as relevant to phase III trials conducted in other fields of medicine because it was derived and developed exclusively from oncology trials. Lastly, although this tool allows any user to obtain inferences from published trial data, we encourage consideration of these limitations and the fact that inference and decision-making are distinct.^26,32^ Clinical trial biostatisticians and principal investigators are the best equipped to select priors and compute probabilities for their specific trial based on the complete underlying individual patient-level data and content-matter expertise in their field.

In summary, the present study provides an evidence-based off-the-shelf prior that can be used to improve trial interpretation by enabling computation of the probabilities of any benefit and a clinically meaningful benefit based on summary statistics from published Cox regression analyses. This tool is freely available online, without the need for coding. Practicing oncologists, patients, scientists, and students may find estimation of posterior probabilities to be valuable for placing trial results in context. We encourage clinical trial principal investigators, regulators, and other stakeholders to consider computing and reporting the probability of whether a treatment is provides a clinically meaningful benefit, and not just the *P* value, when weighing the merits and drawbacks of a new treatment.

## Supporting information

Supplemental, prior derivation

Dataset, summary statistics

## Data Availability

The underlying dataset, code for the development of the prior, and code for computing posterior probability are provided in the Supplement. A freely available webpage has been created for users to compute the probabilities at a given level of hazard ratio based on trial summary statistics.

https://alexandersherry.shinyapps.io/shinyapp/

## Acknowledgments

We thank Erica Goodoff, Senior Scientific Editor in the Research Medical Library at The University of Texas MD Anderson Cancer Center, for editing this article.

