## Supplemental tables and figures for "An Evidenced-Based Prior for Estimating the Treatment Effect of Phase III Randomized Trials in Oncology"

**Figure S1**. Trial selection and flowchart.

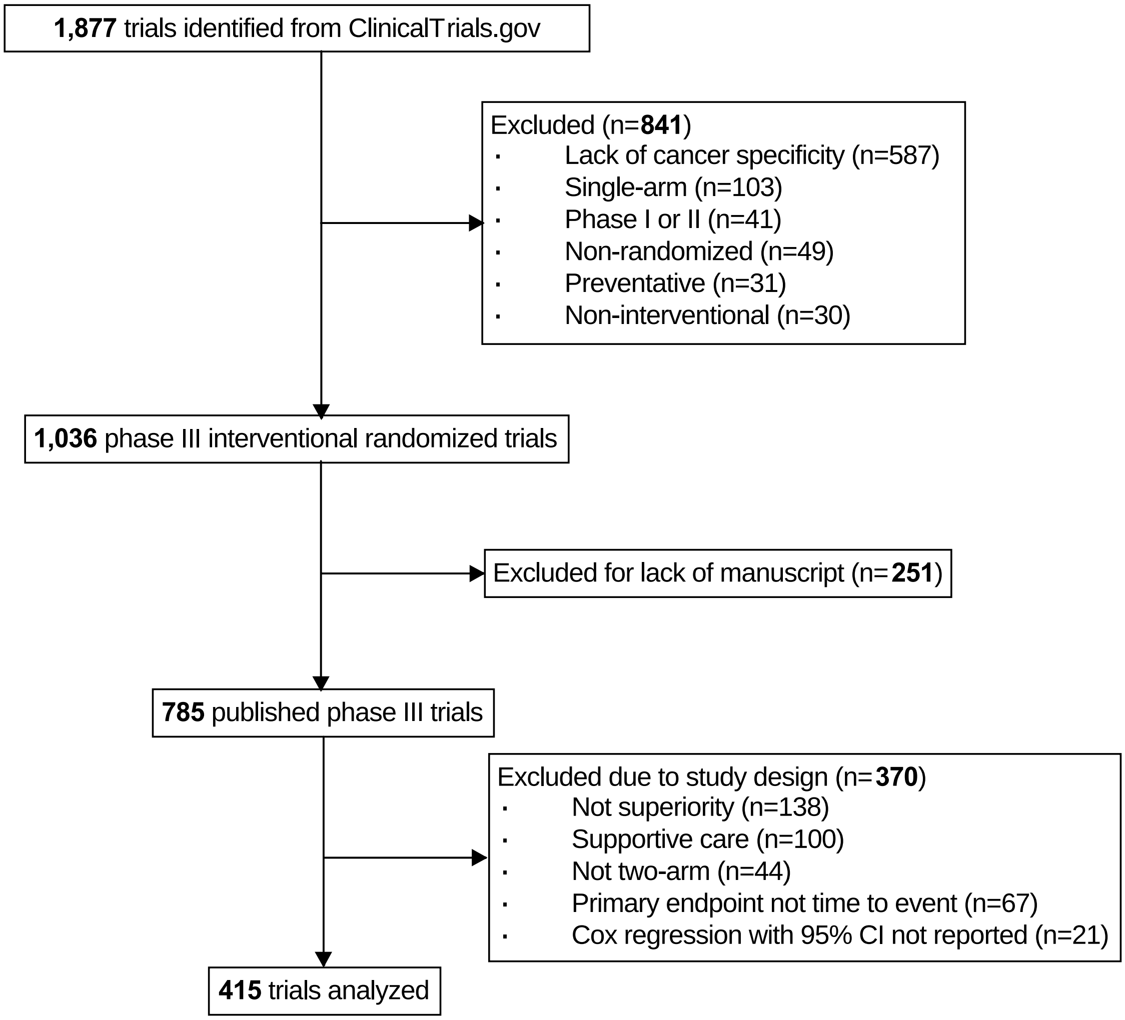

**Figure S2.** Comparison of the absolute z-statistics between RCTs from the Cochrane database of systematic reviews (CDSR) to phase III oncology trials. Violin plots are shown with boxplots representing the median and interquartile range. The absolute z-statistics of the phase III oncology trials were larger than the RCTs from the CDSR, *P* < 0.0001 by Wilcoxon rank sum test.

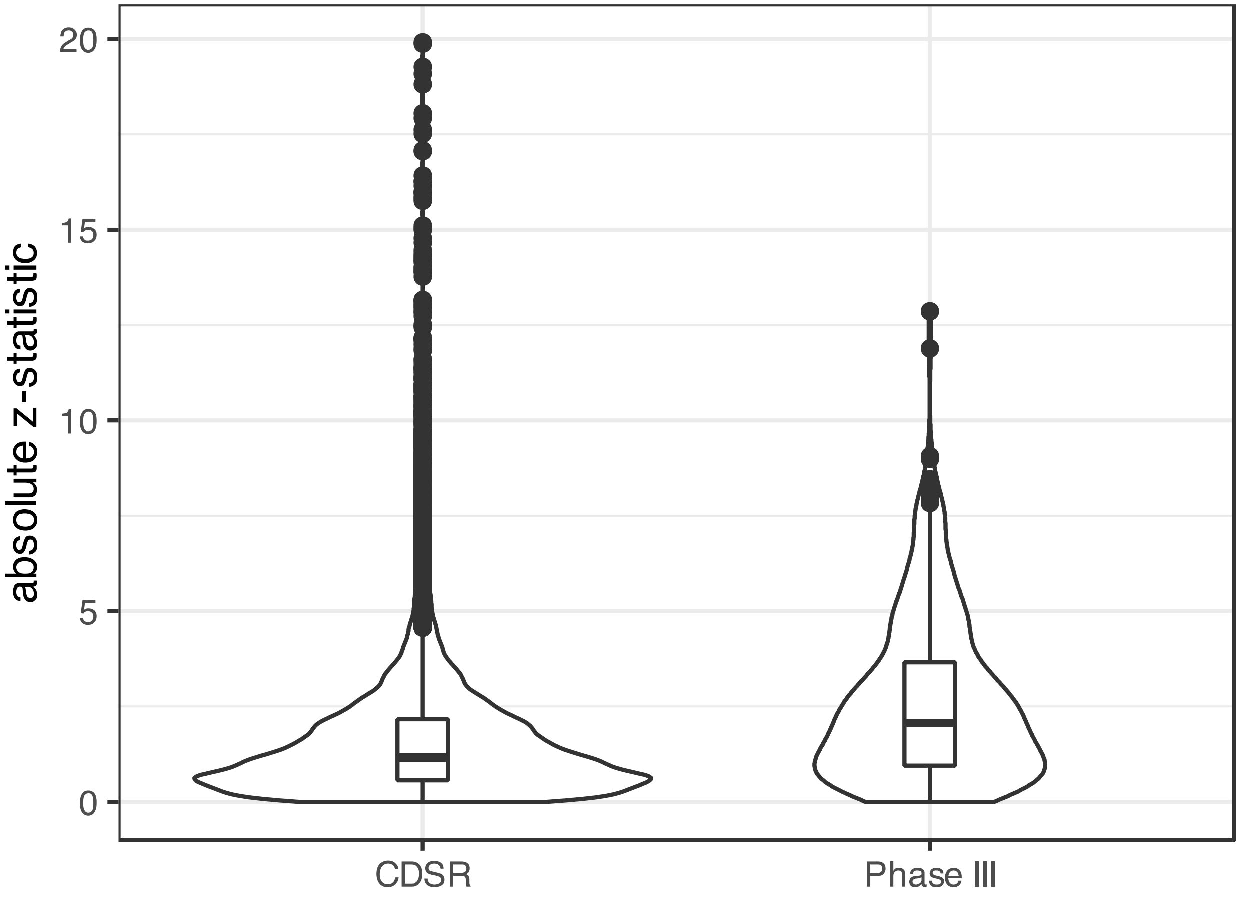

**Figure S3.** Fit of the prior mixture against the absolute z-scores of phase III oncology trials.

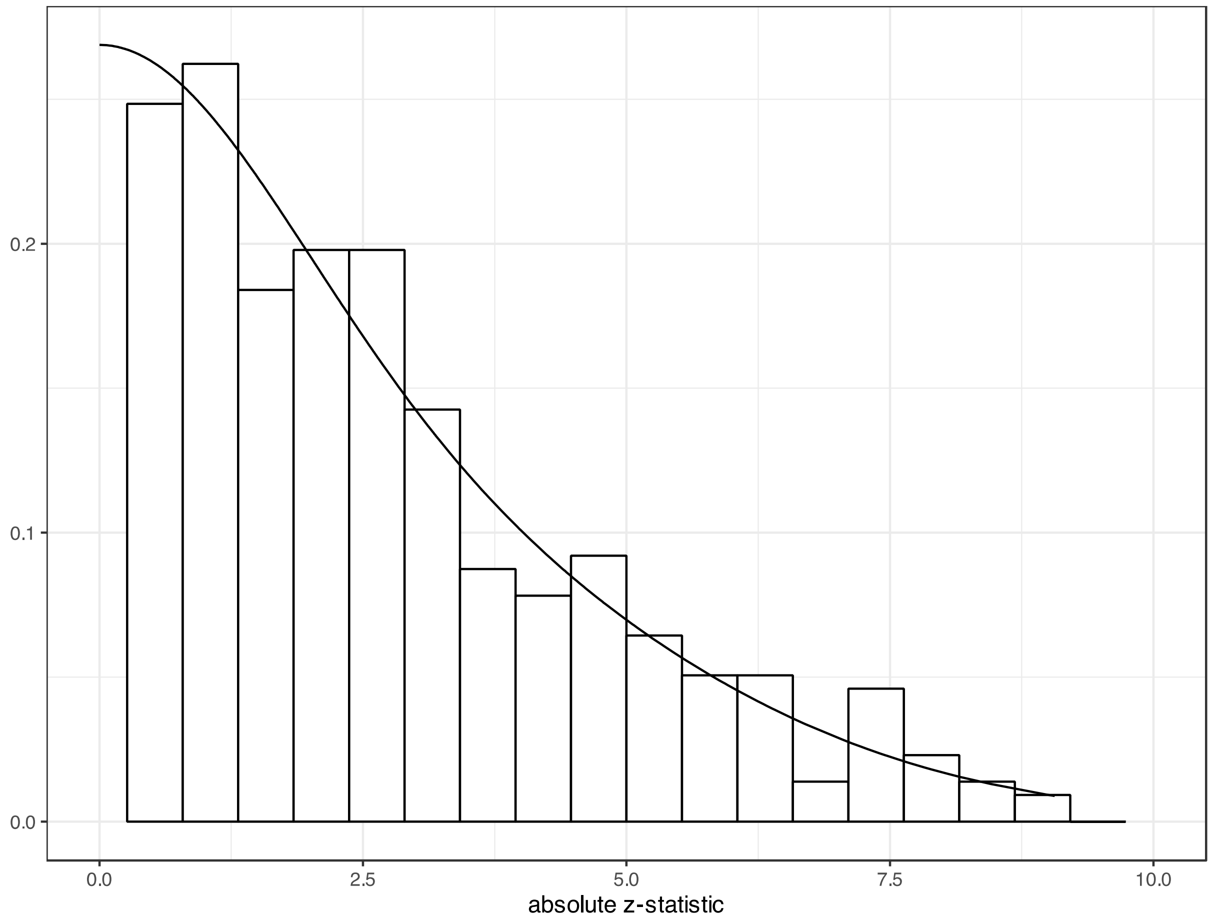

**Figure S4**. The phase III oncology prior is based on mixture distributions, and so is not represented by a single distribution curve. Because the SNR distribution was scaled by the observed standard error for each trial, there are 415 different probability density curves representing the phase III oncology-specific prior, which are displayed here.

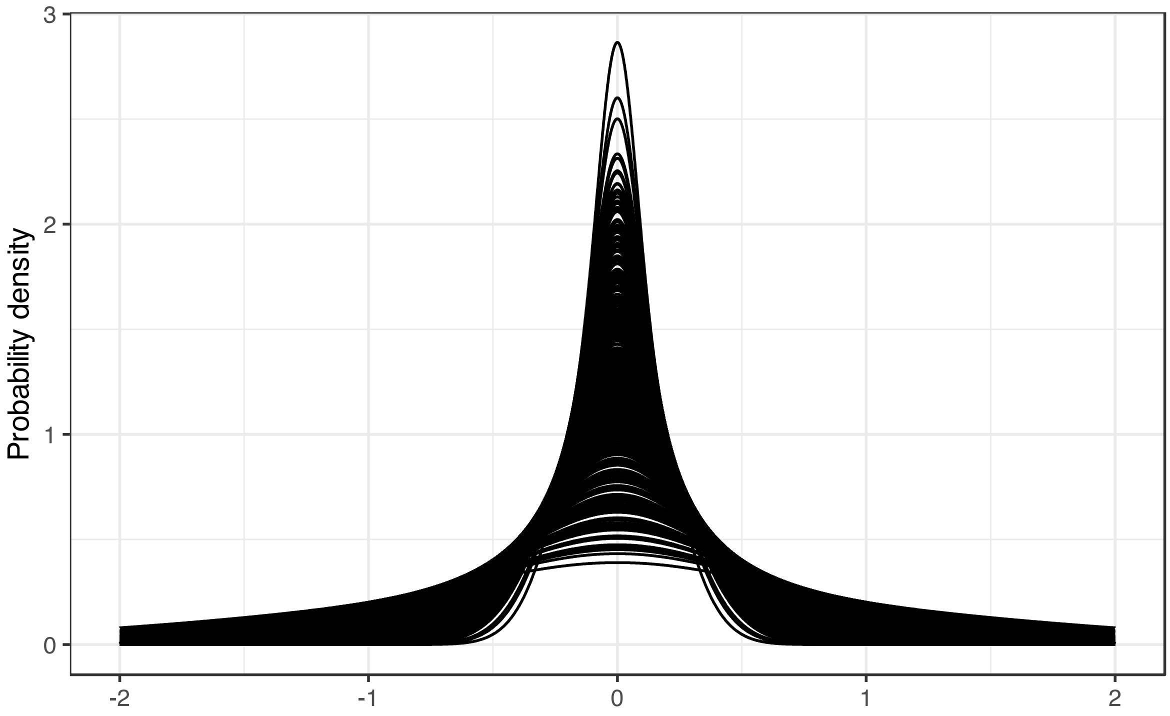

**Figure S5.** Representative examples of probability density distributions from the mixture, including two extreme distributions and one intermediate distribution.

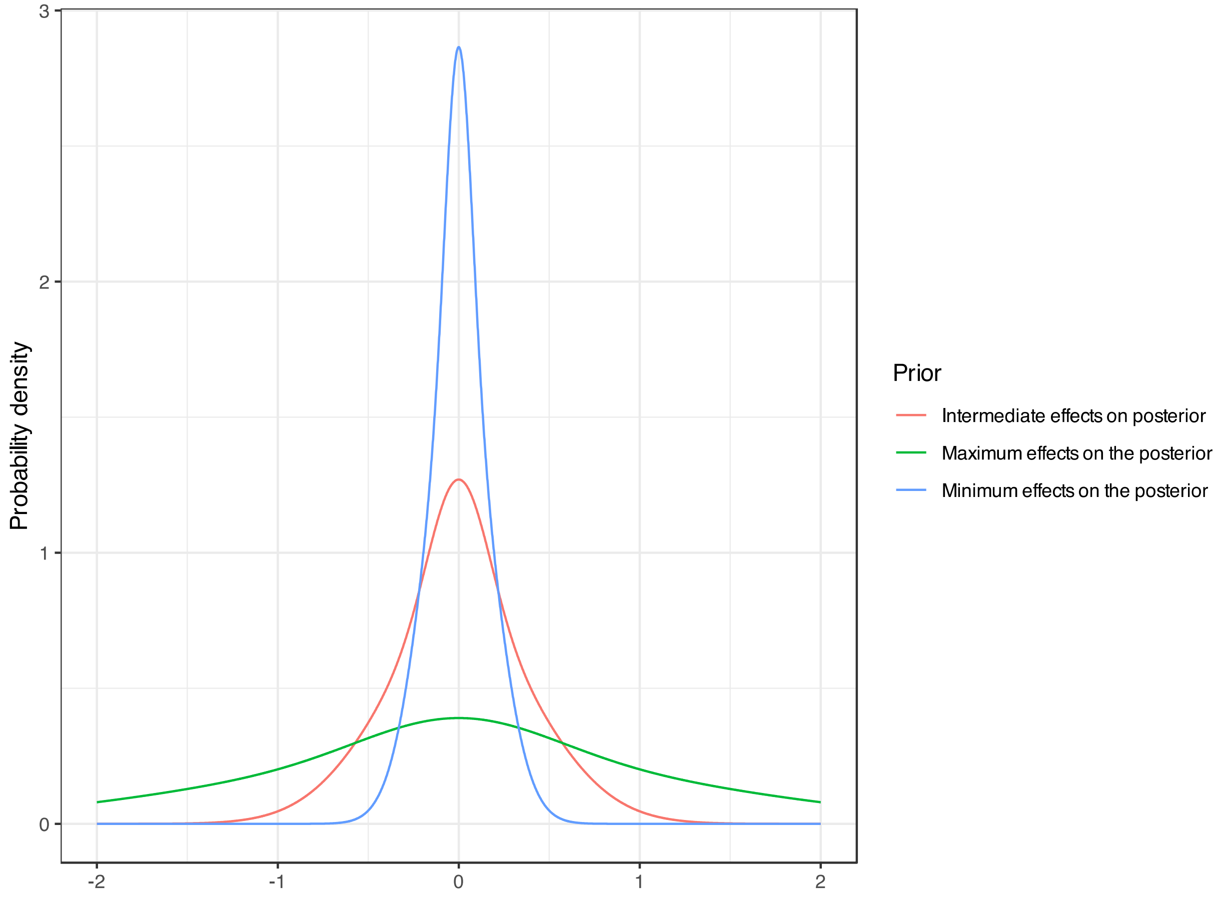

**Table S1**. Components of the phase III oncology-specific mixture prior. Abbreviations: SD, standard deviation; SNR, signal-to-noise ratio.

| | **Component** | | |
|---|---|---|---|
| **Property** | **1** | **2** | **3** |
| Proportion | 0.01373314 | 0.20149644 | 0.78477042 |
| Mean | 0 | 0 | 0 |
| SD z statistic | 1.685708 | 1.694712 | 3.739602 |
| SD SNR | 1.35706 | 1.368228 | 3.603418 |
