## Supplementary material for "An Evidenced-Based Prior for Estimating the Treatment Effect of Phase III Randomized Trials in Oncology": Tables

### **Table 1**. Characteristics of trials included in the analysis.

| **Characteristic** | **No., (%)** |
| --- | --- |
| **Total trials** | 415 |
| Disease stage |  |
| Solid, non-metastatic | 87 (21) |
| Solid, metastatic | 263 (63) |
| Hematologic | 65 (16) |
| Disease site |  |
| Breast | 76 (18) |
| Gastrointestinal | 66 (16) |
| Genitourinary | 58 (14) |
| Hematologic | 65 (16) |
| Thoracic | 79 (19) |
| Other* | 71 (17) |
| Treatment modality |  |
| Systemic therapy | 404 (97) |
| Local therapy | 11 (3) |
| Cooperative group study | 88 (21) |
| Industry sponsored | 361 (87) |
| Median number of enrolled patients (interquartile range) | 596 (377 to 903) |
| Median publication year (interquartile range) | 2015 (2012 to 2017) |
| Primary endpoint |  |
| Overall survival | 165 (40) |
| Surrogate | 250 (60) |
| Primary outcome |  |
| Superiority shown for experimental arm | 216 (52) |
| Superiority not shown for experimental arm | 191 (46) |
| Inferiority of experimental arm | 8 (2) |

*Other disease sites included: central nervous system, endocrine, gynecologic, head and neck, pediatric, sarcoma, and skin.

**Table 2**. Probabilities of benefit (hazard ratio [HR] < 1) and achieving a minimum clinically important difference (MCID) (HR < 0.8) in phase III oncology randomized clinical trials, computed by a phase III, oncology-specific prior.

| **Posterior probability** | **RCTs grouped by trial result interpretation, no. (%)** | |
| --- | --- | --- |
|  | Superiority claimed for the experimental arm, n=216 | Superiority not claimed for the experimental arm, n=199 |
| MCID (HR < 0.8) |  |  |
| ≥ 90% | 114 (53) | 0 (0) |
| ≥ 75% | 149 (69) | 1 (0.5) |
| ≥ 50% | 180 (83) | 6 (3) |
| Any benefit (HR < 1) |  |  |
| ≥ 90% | 216 (100) | 34 (17) |
| ≥ 75% | 216 (100) | 82 (41) |
| ≥ 50% | 216 (100) | 130 (65) |
