## Supplemental, prior derivation for "An Evidenced-Based Prior for Estimating the Treatment Effect of Phase III Randomized Trials in Oncology"

#### Contents


**1 Packages** 1

**2 Key reference** 1

**3 Read the data** 2

**4 Functions** 2

**5 Priors** 3

**6 Phase III oncology prior** 3

**7 Fit of the Phase III prior** 5

**8 Comparing the SNR of Phase III trials to randomized clinical trials (RCTs) in the Cochrane database** 6

**9 Calculate posterior probabilities of the minimum clinically important difference (MCID) and any benefit** 7

**10 Power** 7


#### 1 Packages

```
suppressPackageStartupMessages({
  library(readxl)
  library(dplyr)
  library(ggplot2)
})
```

#### 2 Key reference

van Zwet E, Schwab S, Senn S. The statistical properties of RCTs and a proposal for shrinkage. Statistics in Medicine. 2021;40(27):6107-6117. doi:https://doi.org/10.1002/sim.9173

### 3 Read the data

```
d=read_xlsx("~/Desktop/Summary_statistics.xlsx")
d$b=d$lnHR
d=rename(d,se=s)
d$pval=2*pnorm(-abs(d$z))
```

### 4 Functions

```
dmix = function(x,p,m,s){ # density of normal mixture (vector x)
  drop(p %*% sapply(x, function(x) dnorm(x,mean=m,sd=s)))
}

pmix = function(x,p,m,s){ # cdf of normal mixture (vector x)
  drop(p %*% sapply(x, function(x) pnorm(x,mean=m,sd=s)))
}

rmix = function(n,p,m,s){ # density of normal mixture
  d=rmultinom(n,1,p)
  rnorm(n,m%*%d,s%*%d)
}

# minus log likelihood
loglik = function(theta,z,k){
  p=c(theta[1:(k-1)],1-sum(theta[1:(k-1)]))
  s=theta[k:(2*k-1)]
  m=rep(0,k)
  lik=dmix(z,p,m=m,s=s)
  return(-sum(log(lik)))   # *minus* the log lik
}

# Estimate k-component zero-mean normal mixture distribution
# Uses base R function `constrOptim` to run constrained optimization.
# the constraints such that the mixture proportions are non-negative
# and add up to 1, and the component variances are at least 1.

mix = function(z,k=3){
  # set up constraints for optimization
  # The feasible region is defined by ui %*% par - ci >= 0
  ui=c(rep(-1,(k-1)),rep(0,k))         # (k-1) mixture props sum to < 1
  ui=rbind(ui,cbind(diag(2*k-1)))
  ci=c(-1,rep(0,k-1),rep(1,k))

  # set starting value
  theta0=c(rep(1/k,(k-1)),c(1.2,2:k))
  opt=constrOptim(theta=theta0,f=loglik,ui=ui,ci=ci,
                  method = "Nelder-Mead", z=z,k=k,
                  control=list(maxit=10^4))

  # collect the results
```

```r
  p=c(opt$par[1:(k-1)],1-sum(opt$par[1:(k-1)]))  # mixture proportions
  sigma=opt$par[k:(2*k-1)]                        # mixture sds
  m=rep(0,k)                                      # mixture means
  df=data.frame(p=p,m=m,sigma=sigma)
  return(df)
}


# p(beta | b,se) when beta ~ dmix(p,m,s)
posterior = function(b,se,p,m,s) {
  se2 = se^2
  s2 = s^2
  p = p*dnorm(b,m,sqrt(s2+se2))
  p = p/sum(p)                       # conditional mixing probs
  m = b*s2/(s2+se2) + m*se2/(s2+se2) # conditional means
  v = s2*se2/(s2+se2)                # conditional variances
  s = sqrt(v)                        # conditional std devs
  data.frame(p,m,v,s)
}

# compute posterior mean and two posterior probs
inference = function(b,se,p,m,s){
  post=posterior(b,se,p,m,s)
  pm=sum(post$p*post$m)
  p1=pmix(log(1),p=post$p,m=post$m,s=post$s)
  p08=pmix(log(0.8),p=post$p,m=post$m,s=post$s)
  data.frame(pm,p1,p08)
}
```

### 5 Priors

The zero-mean normal mixture distribution from van Zwet, Schwab and Senn (2021) is defined by:

```r
m=rep(0,4)
p=c(0.32,0.31,0.3,0.07)
sigma=c(1.17,1.74,2.38,5.73)
s= sqrt(sigma^2 - 1)
```

### 6 Phase III oncology prior

We fit a 3-component zero-mean normal mixture to 415 phase III oncology studies

```r
dist=mix(d$z,k=3)

P=dist$p
M=dist$m
Sigma=dist$sigma
S=sqrt(Sigma^2 - 1)
dist
```

```
           p m    sigma
1 0.01373314 0 1.685708
2 0.20149644 0 1.694712
3 0.78477042 0 3.739602
```

This mixture distribution can be used as a prior for the signal-to-noise ratio (SNR). To use it as a prior for the treatment effect, it must be scaled by the observed standard error. So, there is not a *single* prior. To illustrate, we show two extreme cases and an intermediate case.

```
x=seq(-2,2,0.001)
prior_min=data.frame(x,prior=dmix(x,p=P,m=M,s=min(d$se)*S),
                     Prior="Minimum effects on the posterior")
prior_median=data.frame(x,prior=dmix(x,p=P,m=M,s=median(d$se)*S),
                        Prior="Intermediate effects on posterior")
prior_max=data.frame(x,prior=dmix(x,p=P,m=M,s=max(d$se)*S),
                     Prior="Maximum effects on the posterior")
prior=rbind(prior_min,prior_median,prior_max)
ggplot(prior,aes(x=x,y=prior,group=Prior,color=Prior)) +
  geom_line() + xlab("") +ylab("Probability density") + theme_bw()
```

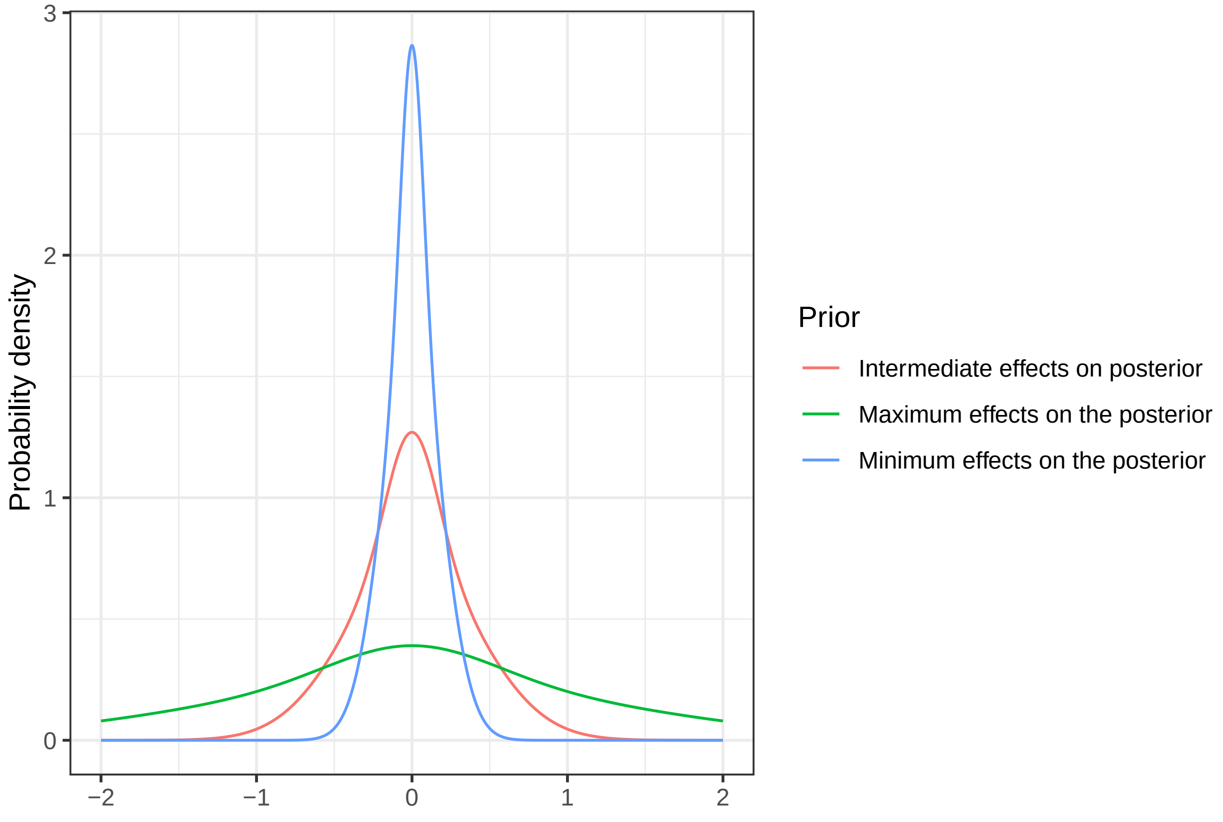

#### 7 Fit of the Phase III prior

The fit of this mixture distribution to the z values of the 415 phase III oncology trials is quite reasonable.

```
d$fz=dmix(abs(d$z),p=P,m=M,s=Sigma)
ggplot(d, aes(x = abs(z))) +
  geom_histogram(aes(y = after_stat(density)), color="black",
                 fill="white", bins=20) +
  geom_line(aes(x=abs(z),y=2*fz)) +
  xlim(0,10) + xlab("absolute z-statistic") + ylab('') + theme_bw()
```

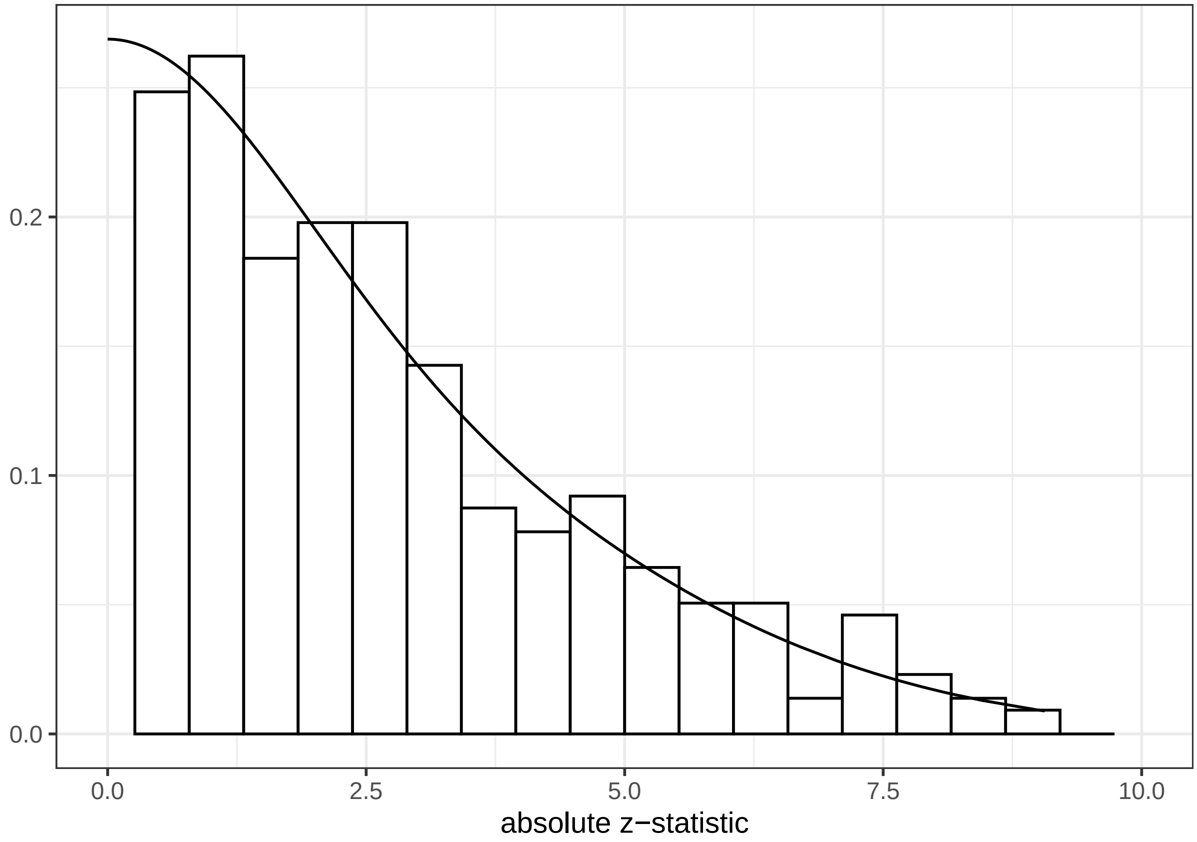

### 8 Comparing the SNR of Phase III trials to randomized clinical trials (RCTs) in the Cochrane database

If we compare to the distribution from van Zwet, Schwab and Senn(2021), we see that the SNR of a phase III study tends to be much larger than a typical trial from the Cochrane database.

```
x=seq(-10,10,0.001)
prior1=data.frame(x,prior=dmix(x,p=p,m=m,s=s),
                  Source="23,551 RCTs Across the Cochrane Database")
prior2=data.frame(x,prior=dmix(x,p=P,m=M,s=S),
                  Source="415 Phase III Oncology RCTs")
prior=rbind(prior1,prior2)
ggplot(prior,aes(x=x,y=prior,group=Source,color=Source)) +
  geom_line() + ylab("Density of SNR Distribution") + xlab("SNR") + theme_bw() +
  ggtitle("RCT Signal-to-Noise Ratios Across Medicine vs Phase III Oncology") +
  theme(plot.title = element_text(hjust = 0.5, size =13),
        legend.text = element_text(size = 11)) +
  theme(legend.position = "top") + guides(color = guide_legend(title = NULL))
```

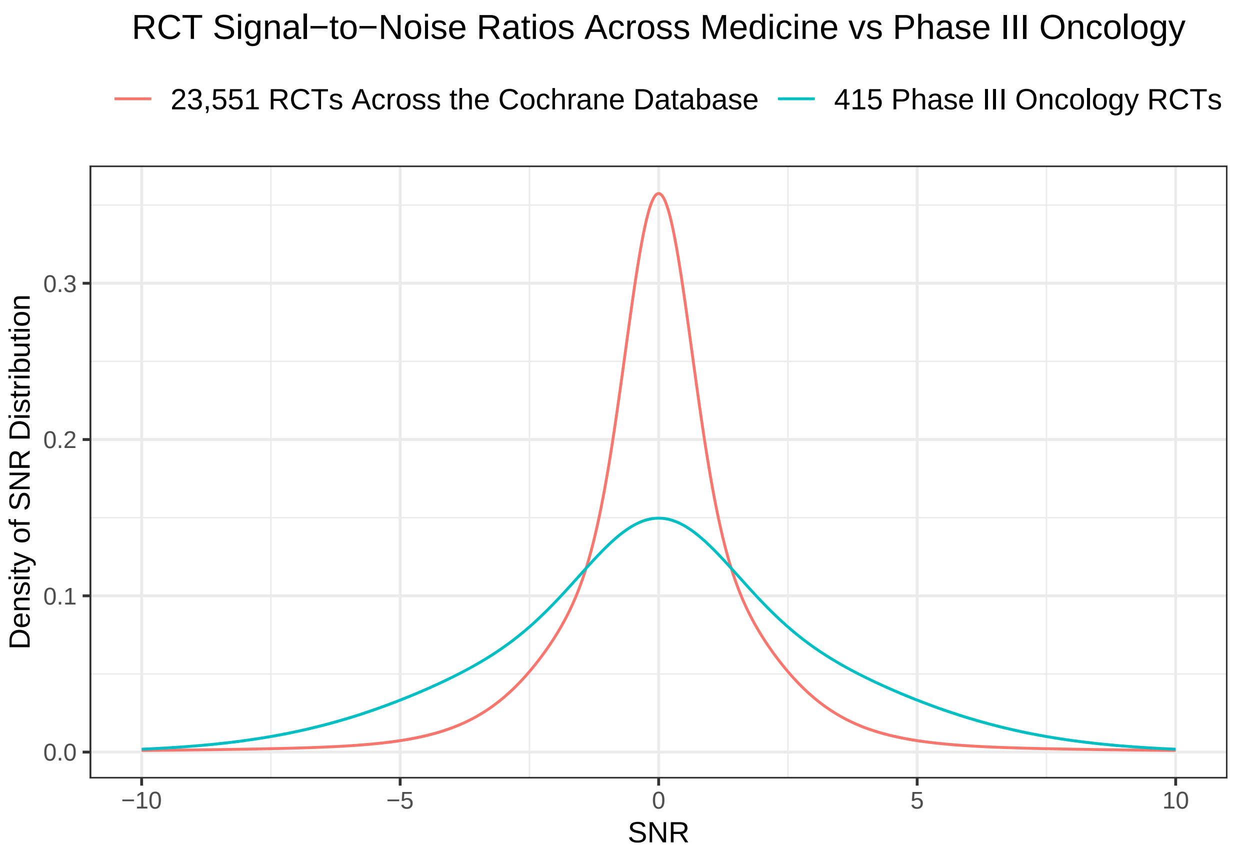

### 9 Calculate posterior probabilities of the minimum clinically important difference (MCID) and any benefit

We compute the posterior mean and the posterior probabilities of any benefit (HR < 1) and the MCID (HR < 0.8) for each of the 415 RCTs.

```
df1=d %>% rowwise %>% mutate(inference(b,se,p=P,m=M,s=S*se))
```

### 10 Power

We can transform the estimated distribution of the SNR among phase III trials in oncology into the distribution of their power.

```
snr=rmix(10^6,p=P,m=M,s=S)
power=pnorm(-1.96,snr,1) + 1 - pnorm(1.96,snr,1)
summary(power)
```

```
   Min. 1st Qu.  Median    Mean 3rd Qu.    Max. 
 0.0500  0.1379  0.4854  0.5239  0.9505  1.0000 
```

```
df=data.frame(snr,power)
ggplot(df, aes(x = power)) +
  geom_histogram(aes(y = after_stat(density)), color="black",
                 fill="white", bins=100) +
  xlab("power") + ylab('') + theme_bw() +
  ggtitle("Observed Power in Phase III Oncology RCTs")+
  theme(plot.title = element_text(hjust = 0.5, size =15))
```

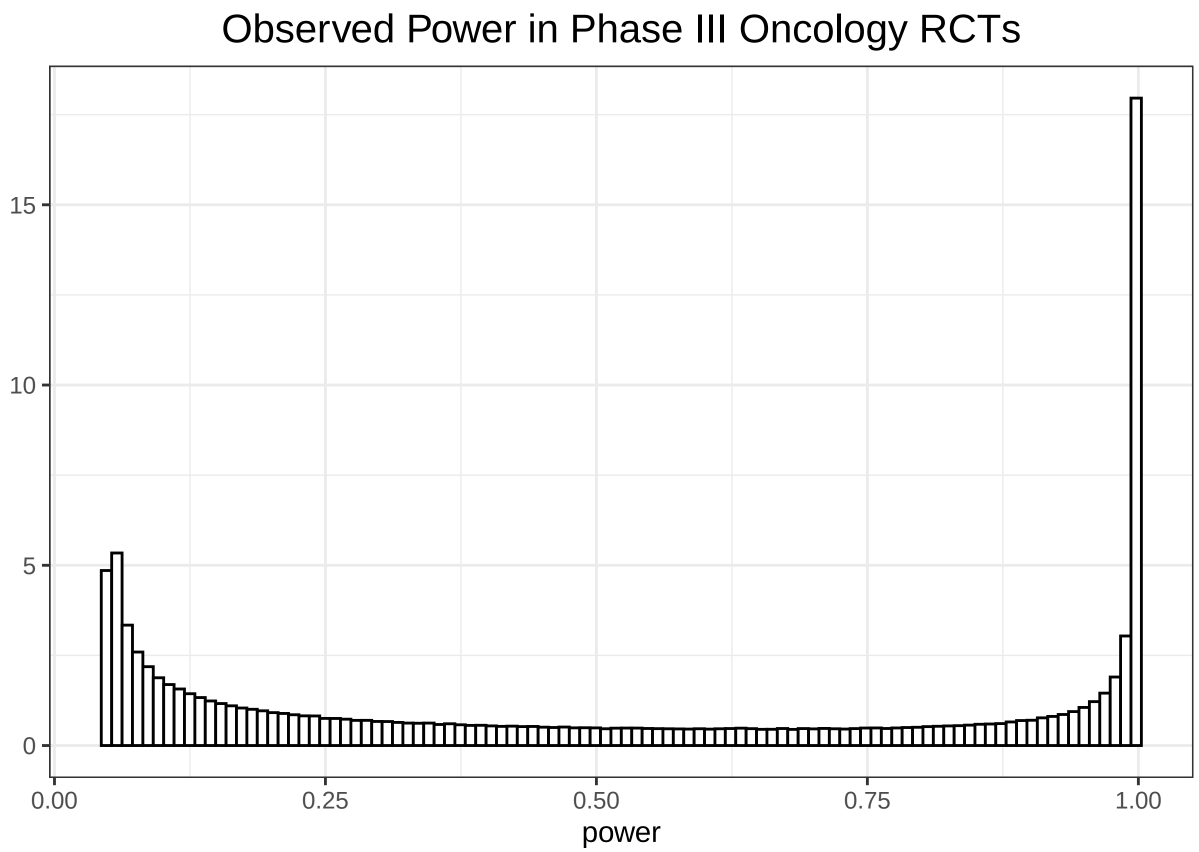

Observed Power in Phase III Oncology RCTs
15
10
5
0
0.00
0.25
0.50
0.75
1.00
power
